# Rationale, Design, and Baseline Characteristics of the VALIANT (COVID-19 in Older Adults: A Longitudinal Assessment) Cohort

**DOI:** 10.1101/2022.09.14.22279932

**Authors:** Andrew B. Cohen, Gail J. McAvay, Mary Geda, Sumon Chattopadhyay, Seohyuk Lee, Denise Acampora, Katy Araujo, Peter Charpentier, Thomas M. Gill, Alexandra M. Hajduk, Lauren E. Ferrante

## Abstract

**Background:** Most older adults hospitalized with COVID-19 survive their acute illness. The impact of COVID-19 hospitalization on patient-centered outcomes, such as physical function, cognitive function, and symptoms, is not well understood. We sought to address this knowledge gap by collecting longitudinal data about these issues from a cohort of older adult survivors of COVID-19 hospitalization.

**Methods:** We undertook a prospective study of community-living persons age ≥60 years who were hospitalized with COVID-19 from June 2020 to June 2021. A baseline interview was conducted during or up to two weeks after hospitalization. Follow-up interviews occurred at one, three, and six months post-discharge. In interviews, participants completed comprehensive assessments of physical and cognitive function, symptoms, and psychosocial factors. If a participant was too impaired to complete an interview, an abbreviated assessment was performed with a proxy. Additional information was collected from the electronic health record. Baseline characteristics of the cohort are reported here.

**Results:** Among 341 participants, the mean age was 71.4 (SD 8.4) years, 51% were women, and 37% were of Black race or Hispanic ethnicity. Median length of hospitalization was 8 (IQR 6-12) days. All but 4% of participants required supplemental oxygen and 21% required a higher level of care in an intensive care unit or stepdown unit. Nearly half (47%) reported at least one disability in physical function, 45% demonstrated cognitive impairment, and 67% were pre-frail or frail. Participants reported a mean of 9 of 14 (SD 3) COVID-19-related symptoms.

**Conclusions:** Older adults hospitalized with COVID-19 demonstrated high rates of baseline physical and cognitive impairment as well as high symptom burden. Longitudinal findings from this cohort will advance our understanding of outcome trajectories of great importance to older survivors of COVID-19.

## Introduction

Older adults are much more likely than younger adults to experience serious illness with COVID-19. A study in New York City during the first wave of infection found that the odds of hospitalization for persons ≥75 years were almost 40 times higher than for those under age 65.^1^ The most recent data from the Centers for Disease Control and Prevention, reported on June 2, 2022, show that 74% of deaths — more than 748,000 — have occurred among Americans age 65 and older.^2^ This amounts to more than 1 in 100 older Americans.

Nonetheless, most older adults survive serious illness (i.e., illness requiring hospitalization) with COVID-19.^3^ There is reason to be concerned, however, that older survivors are at risk for substantial decline in their health and functional status and that they may develop impairments that last even after the pandemic wanes. Normal aging confers decreased physiologic reserve and diminished resilience in the face of acute stressors,^4^ and older adults are much more likely than younger adults to have multiple chronic medical conditions as well as other underlying vulnerabilities, such as frailty.^5^ Hospitalization with COVID-19 involves long periods of isolation from all but essential medical staff, during which patients are unable to leave their hospital rooms, are at increased risk for immobility,^6^ and experience profound social isolation.^7^ Delirium is also common.^8^ Upon discharge, there may be limited access to physical therapy and other rehabilitative services.^9^

There are few data from those who have been hospitalized with COVID-19 about the health outcomes that matter most to older patients, including preservation of cognition, maintaining physical function, and freedom from burdensome symptoms.^10^ This is in part because information about these issues is not collected in routine clinical care. To address this knowledge gap, we designed VALIANT (CO<u>V</u>ID-19 in Older Adults: <u>A L</u>ongitudinal <u>A</u>ssessme<u>nt</u>), a prospective longitudinal study of adults ≥ 60 years hospitalized with COVID-19 at five hospitals in Connecticut. We interviewed patients during or shortly after their hospitalization and then at one, three, and six months after hospital discharge in order to collect granular information across multiple health domains. We then linked these data to information from the electronic medical record. The resulting dataset will permit an unusually rich examination of outcomes and outcome trajectories among older adults who require hospitalization for COVID-19.

## Methods

### Study Overview

VALIANT is a prospective longitudinal study of community-living persons age ≥60 years who were hospitalized with COVID-19 at one of five hospitals within the Yale-New Haven Health System (YNHHS) from June 2020 to June 2021. During a baseline interview and assessment, conducted during hospital admission or within two weeks of discharge, we collected information about participants’ physical function, cognitive function, symptoms, and psychological and lifestyle factors. Data about the same issues were subsequently collected at follow-up interviews at one, three, and six months after discharge. All assessments occurred via phone or videoconferencing software. Patient-reported data were supplemented by data from the electronic health record. The study was approved by the Institutional Review Board at Yale University. Methods reporting is consistent with the STROBE guidelines.^11^

### Patient Screening and Eligibility

Trained research coordinators used the electronic health record to identify potential participants. Patients were eligible if they were at least 60 years old, hospitalized at a YNHHS hospital, and had a PCR-confirmed diagnosis of SARS-CoV-2 infection either during or directly prior to hospital admission. Patients were excluded if they had previously been hospitalized with COVID-19 and were subsequently readmitted, regardless of whether their SARS-CoV-2 test remained positive. We made an exception for patients who were readmitted within 7 days of hospital discharge from their initial (index) COVID-19 admission, because, in such cases, we considered the two hospitalizations to constitute a single episode of care. Patients were also excluded if they were long-term residents in a skilled nursing facility, had advanced dementia, did not speak English or Spanish, or had opted out of research. Additionally, because we wished to collect longitudinal data about persons who were not at the end of life, patients were excluded if they had “comfort measures only” orders or had a planned discharge to hospice. The charts of all patients enrolled in the study were reviewed by one of the three principal investigators (A.B.C., a geriatrician; A.M.H., an epidemiologist with training in aging research; and L.E.F., a critical care physician) to confirm their eligibility.

For potential participants for whom there was concern about decisional impairment, we conducted the University of California San Diego Brief Assessment for Capacity to Consent.^12^ If decisional impairment was confirmed, proxies were sought for the informed consent process, along with verbal assent from participants. Proxies were also sought for participants who were unable to consent due to severity of illness (e.g., those who were intubated).

Participants or proxies were contacted via phone or videoconferencing software by research coordinators who explained the study, answered questions, and completed a verbal informed consent process. Consent information sheets were mailed to the patient’s home.

### Assessment Schedule

All study assessments were completed via phone or videoconference for the convenience of participants and safety of the study team. A schedule of the study interviews appears in **Figure 1**. Participants underwent a baseline interview, conducted in English or Spanish, during their COVID-19 hospitalization or within two weeks of hospital discharge. Completion of the baseline interview was required for the participant to be considered enrolled in the study. We collected information about pre-hospitalization physical function, health status, symptoms, and psychological and lifestyle factors. We also performed cognitive testing, described in detail below. Proxies completed an abbreviated interview that omitted cognitive testing as well as certain measures that could only be reliably reported by patients, such as depressive and anxiety symptoms. Follow-up interviews containing the same or similar measures occurred at one, three, and six months from the date of hospital discharge or, for readmitted patients, from date of discharge from the readmission. Follow-up interviews were completed by patients whenever possible or by proxy. A summary of assessments at each time point appears in **Supplemental Table 1** and further details for select assessments appear in **Supplemental Table 2**.

**Figure 1.**
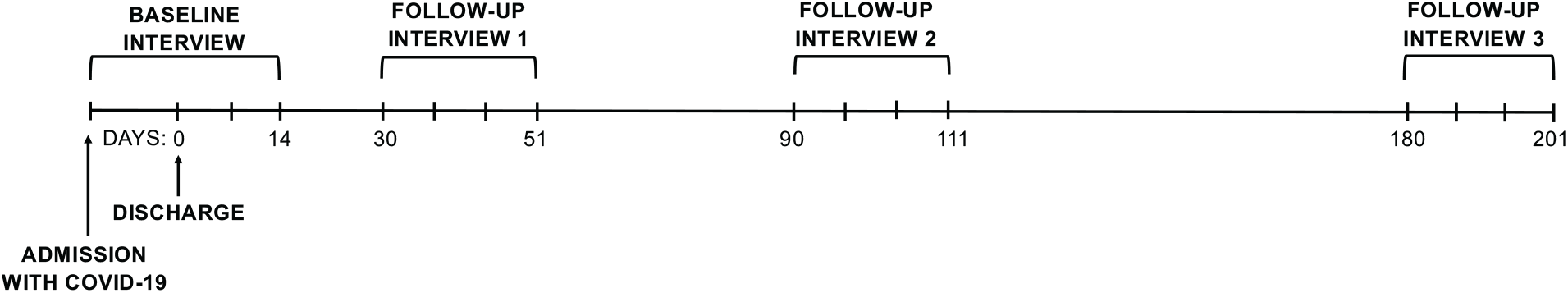
Schedule of study assessments for VALIANT. The baseline interview occurred during hospitalization with COVID-19 or within 2 weeks of discharge. For patients who were discharged and readmitted within 7 days, the baseline interview could occur within 2 weeks of discharge from the readmission. Follow-up interviews occurred during 3-week windows that opened at 1 month (30 days), 3 months (90 days), and 6 months (180 days) after discharge. Each tick mark (“|”) in the figure represents one week.

### Primary Measures

#### Physical Function

Physical function was assessed using a 15-item scale that captures disability in seven basic activities of daily living (bathing, dressing, transferring from a chair, walking inside the house, eating, toileting, and grooming),^13^ five instrumental activities of daily living (shopping, housework, meal preparation, taking medications, and managing finances),^14^ and three mobility activities (walking a quarter mile, climbing a flight of stairs, and lifting or carrying ten pounds).^15^ During the baseline interview, participants were asked about disability in these 15 functional activities at one month prior to admission to evaluate their pre-illness status. Disability in each functional activity was defined as needing help from another person to complete the task. In follow-up assessments at one, three, and six months post-discharge, participants were asked about disability in the same 15 functional activities “at the present time.”

#### Cognitive Function

Cognition was assessed using the Montreal Cognitive Assessment (MoCA) 5-minute protocol,^16^ an abbreviated version of the MoCA that has been modified for administration by telephone. The MoCA 5-minute protocol measures attention, orientation, language, verbal learning and memory, and executive function. It displays psychometric properties similar to the full MoCA^17^ and utilizes a cutoff <22 (range, 0 to 30) to indicate cognitive impairment.

#### Symptoms

When data collection commenced in June 2020, a symptom assessment instrument specific to COVID-19 had not yet been developed. We therefore adapted an existing instrument commonly used to measure symptoms in older adults, the Edmonton Symptom Assessment System (ESAS).^18^ The ESAS measures the presence and severity of nine common symptoms, including fatigue, pain, and dyspnea. We added symptoms relevant to COVID-19, including cough, anosmia, and ageusia, to comprise a list of 14 symptoms. Given work suggesting that there is considerable variation in the intensity of symptoms among older persons,^19^ we also measured symptom severity, with “1” indicating that a symptom was mild, “2” indicating that it was moderate, and “3” indicating that it was severe, yielding a total symptom score ranging from 0 to 42.

### Secondary Measures

#### Health Status

Health status was assessed with selected questions from the Short Form-12.^20^ As with physical function, participants were asked to report on their health status at one month prior to admission during the baseline interview and on their health status at the present time during follow-up interviews.

#### Frailty

Frailty was assessed in accordance with the Fried phenotype,^21^ with two questions adapted for remote assessment (self-reported walking speed in lieu of a gait test and difficulty opening a jar in lieu of a grip strength test^22^) and the modified questions about low physical activity used in the National Health and Aging Trends Study.^23^ Frailty was assessed during the baseline and six-month interviews.

#### Sensory Impairments

Vision and hearing were measured using questions from the National Health and Aging Trends Study concerning self-reported impairments in function with or without use of aids.^24^ Hearing impairment was assessed at all time points, given its association with functional disability,^25^ whereas vision impairment was assessed only at baseline.

#### Psychosocial Function

Depressive and anxiety symptoms were assessed with the Patient Health Questionnaire (PHQ-4).^26^ Social support was measured with a five-item version of Medical Outcomes Study Social Support Survey.^27^

#### Physical Activity, Falls, and Fractures

Physical activity level, operationalized as frequency of engagement in light, moderate, and vigorous physical activity, was evaluated using the abbreviated Physical Activity Scale for the Elderly.^28^ Fall history was assessed via a single question regarding a history of falls within the past 12 months. Fracture history was assessed by asking patients whether they had broken a bone since turning 50. Fracture risk was evaluated using selected questions from the Fracture Risk Assessment Tool (FRAX)^29^ and the International Osteoporosis Foundation’s One Minute Risk Check.^30^

#### Post-Discharge Recovery and Events

At follow-up interviews, we collected information from patients about hospital readmissions, emergency department visits, receipt of home health care, receipt of physical or occupational therapy, admission to short-term rehabilitation, fractures during the follow-up period, and restricted activity.

### Medical Record Data

Patient-reported data were supplemented with data from the electronic health record. We obtained granular data about the index hospitalization from the Yale Department of Medicine COVID-19 Data Explorer (DOM-CovX)^31^, which contains health record data from all patients admitted to a YNHHS hospital with a positive SARS-CoV-2 test. Data in the DOM-CovX registry include information about use of specific treatments for COVID-19, such as corticosteroids and remdesivir; severity of illness, as determined by the Sequential Organ Failure Assessment (SOFA) score;^32^ need for advanced respiratory support; and detailed laboratory data. The DOM-CovX registry also provided data about demographics, medical comorbidities, and discharge disposition. Missing data from the registry were filled in via manual chart abstraction, when possible. We supplemented the DOM-CovX registry with additional structured data about delirium, measured by the Confusion Assessment Method (CAM)^33^ or CAM-ICU^34^ and collected as part of routine clinical practice by hospital nursing staff, through a separate electronic health record data pull. We performed additional manual chart abstraction to capture other variables, including information about COVID-19 vaccination status at baseline and follow-up. The Area Deprivation Index, a detailed measure of the socioeconomic conditions in a specific geographical area, was calculated based on each participant’s zip code using the Neighborhood Atlas.^35^

### Data Management

Research Electronic Data Capture (REDCap) software was used for all study workflow organization and data management activities.^36^ Specialized features were developed by the REDCap@Yale team (P.C., S.C., K.A.) at the Yale Claude D. Pepper Older Americans Independence Center. Participant workflow was managed using a custom REDCap dashboard, which incorporated key automation and quality control measures to optimize efficiency of study assessments and accuracy of data collection. Data collection forms utilized integrated quality control checks to identify data entry errors and out-of-range values. DOM-CovX registry data were imported into REDCap. An automated routine created SAS datasets for all study reports, quality control checks, and analyses. Missing data were evaluated for all variables and were addressed using multiple imputation, as appropriate. As diagnostic algorithms for COVID-19 in older adults evolved rapidly, the majority of missing data involve laboratory tests that were introduced or omitted from the YNHHS diagnostic order sets over the course of the study.

### Statistical Analysis

Baseline characteristics of VALIANT participants were summarized using means and standard deviations for normally distributed continuous variables, medians and interquartile ranges for ordinal or non-normally distributed continuous variables, and proportions for categorical variables. Analyses were performed using SAS 9.4 (SAS Institute, Cary, NC).

### Data Sharing

Data from VALIANT will be made available to the public, for use upon approval by the principal investigators, at one year from completion of data collection (January 31, 2023).

## Results

Between June 18, 2020 and June 30, 2021, we screened 1,747 older adults at five hospitals. Among them, 939 were deemed eligible, 680 consented, and 341 were enrolled (**Figure 2)**. There were no differences in demographic characteristics between the screened and enrolled participants. Characteristics of the enrolled participants are presented in **Table 1**. The mean age was 71.4 (standard deviation [SD] 8.4) years, 175 (51.3%) were women, and 125 (36.6%) were of Black race or Hispanic ethnicity. Nearly one third of participants had Medicaid as one of their forms of insurance. Most participants lived at home, with 239 (70.5%) reporting that they lived with others and 92 (27.1%) reporting that they lived at home alone.

**Figure 2.**
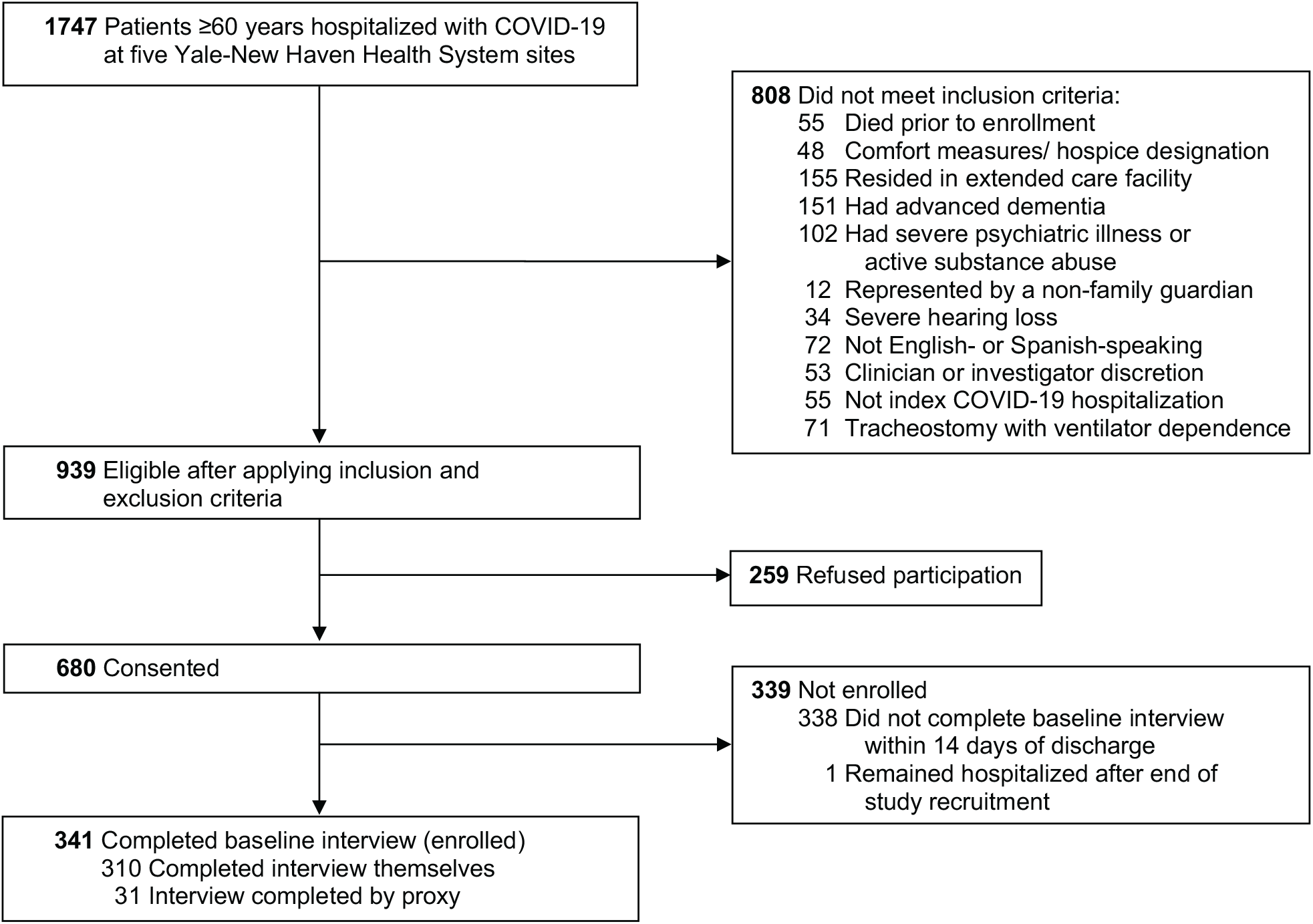
Screening and enrollment flow diagram for participants in VALIANT.

**Table 1.** Sociodemographic characteristics of VALIANT participants

| **Characteristic** | **VALIANT Participants**<br>**(N = 341)<sup>a</sup>** |
| --- | --- |
| Age, years, mean (SD) | 71.4 (8.4) |
| Female sex, n (%) | 175 (51.3) |
| Race and ethnicity, n (%) |  |
| White, non-Hispanic | 211 (61.9) |
| White, Hispanic | 8 (2.3) |
| Black, non-Hispanic | 79 (23.2) |
| Black, Hispanic | 1 (0.3) |
| Hispanic, race not reported | 37 (10.9) |
| Non-Hispanic, race not reported | 1 (0.3) |
| Asian, non-Hispanic | 3 (0.9) |
| Unknown race and ethnicity | 1 (0.3) |
| Primary language, n (%) |  |
| English | 315 (92.4) |
| Spanish | 26 (7.6) |
| Highest level of education completed, n (%) |  |
| Less than high school | 50 (14.9) |
| High school or equivalent | 146 (43.6) |
| 2- or 4- year degree | 100 (29.9) |
| Graduate or post-graduate | 39 (11.6) |
| Medicaid recipient, n (%) | 107 (31.4) |
| Living in socioeconomically disadvantaged area, n (%)<sup>b</sup> | 82 (25) |
| Marital status, n (%) |  |
| Married or living with partner | 160 (47.2) |
| Single | 64 (18.9) |
| Divorced or separated | 52 (15.3) |
| Widowed | 62 (18.3) |
| Other | 1 (0.3) |
| Living situation prior to admission, n (%) |  |
| At home alone | 92 (27.1) |
| At home with others | 239 (70.5) |
| Short-term resident at skilled nursing facility | 5 (1.5) |
| Assisted living facility | 3 (0.9) |
| Respondent, n (%) |  |
| Participant | 309 (90.6) |
| Proxy | 32 (9.4) |

<sup>a</sup> There were missing data for the following variables (number of participants with missing values noted in parentheses): highest level of education completed (6), living in socioeconomically disadvantaged area (13, due to inability to calculate Area Deprivation Index), marital status (2), living situation (2).
<sup>b</sup> Area Deprivation Index score >55

Clinical and functional characteristics of study participants before their hospital admission with COVID-19 are provided in **Table 2**. Participants had a median of three comorbidities (interquartile range [IQR] 1-4), and 157 (46.6%) reported having at least one disability in the month prior to hospitalization. Of the 301 participants with complete frailty data, 56 (18.6%) were frail, 147 (48.8%) were pre-frail, and 98 (32.6%) were not frail. There were 52 participants (15.2%) who had received at least one dose of a COVID-19 vaccine before being hospitalized.

**Table 2.** Clinical characteristics of VALIANT participants prior to COVID-19 hospitalization

| Characteristic | VALIANT Participants<br>(N = 341)<sup>a</sup> |
| --- | --- |
| General health in month prior to hospitalization, n (%) |  |
| Excellent | 47 (15.2) |
| Very good | 89 (28.7) |
| Good | 99 (31.9) |
| Fair | 63 (20.3) |
| Poor | 12 (3.9) |
| Comorbidity count (range, 0-11), median (IQR)<sup>b</sup> | 3 (1-4) |
| Disability in month prior to hospitalization |  |
| Number of functional activities in which participant experienced disability (range, 0-15), median (IQR)<sup>c</sup> | 0 (0-4) |
| Presence of disability in $\geq 1$ functional activity, n (%) | 157 (46.6) |
| Frailty classification in month prior to hospitalization<sup>d</sup> |  |
| Not frail | 98 (32.6) |
| Pre-frail | 147 (48.8) |
| Frail | 56 (18.6) |
| Social support in month prior to hospitalization (range, 5-25), median (IQR)<sup>e</sup> | 23 (20-25) |
| Hearing impairment, n (%)<sup>f</sup> | 120 (35.5) |
| Vision impairment, n (%)<sup>g</sup> | 50 (14.9) |
| Fall in 12 months prior to hospitalization, n (%) | 101 (29.8) |
| History of fracture since age 50, n (%) | 67 (19.8) |
| History of tobacco use | 161 (52.4) |
| $\geq 1$ COVID-19 vaccine dose prior to hospitalization, n (%) | 52 (15.2) |

<sup>a</sup> There were missing data for the following variables (number of participants with missing values noted in parentheses): general health (31), disability in functional activities (4), frailty (40, due to missingness in individual components of the 5-item measure), social support (40), hearing impairment (3), vision impairment (5), history of falls (2), history of fractures (3).
<sup>b</sup> Comorbidities included hypertension, myocardial infarction, heart failure, cerebrovascular disease (stroke, transient ischemic attack, or intracranial hemorrhage), diabetes mellitus, chronic lung disease, chronic kidney disease, end-stage renal disease, liver disease, immunocompromised status (autoimmune disease, HIV positive, or receipt of solid organ transplant), and malignancy (solid tumors, leukemia or lymphoma, or metastatic disease).
<sup>c</sup> Tasks included eating, dressing, bathing, toileting, grooming, getting in and out of a chair, walking around indoors, doing housework, going shopping, preparing a meal, taking medications, managing finances, walking a quarter of a mile, climbing stairs, and lifting or carrying heavy objects. A higher score indicates greater disability.
<sup>d</sup> Frailty was assessed using the Fried frailty score, which consists of five items: low physical activity, slow gait speed, muscle weakness, weight loss, and exhaustion. Participants with scores $\geq 3$ were classified as frail, those with scores of 1-2 were classified as pre-frail, and those with scores of 0 were classified as not frail.
<sup>e</sup> Assessed items included having someone to confide in or talk to about problems, having someone to help with daily chores if experiencing illness, having someone to turn to for suggestions about how to deal with a personal problem, having someone to love and make the participant feel wanted, and having someone to get together with for relaxation or fun. Each item was assessed using a 5-point Likert scale, from 1 (none of the time) to 5 (all of the time). Higher scores indicate greater social support.
<sup>f</sup> Using a hearing aid, not hearing well enough to carry on a conversation in a quiet or loud room, or being deaf.
<sup>g</sup> Having difficulty reading newspaper print, recognizing a person across the street, or watching television from across the room (with use of corrective lenses, when applicable).

Clinical characteristics of study participants during their hospitalization with COVID-19 are shown in **Table 3**. Most were hospitalized on the general wards; however, 40 (11.7%) participants required ICU admission, and 29 (8.5%) required stepdown unit care. Sixty (17.6%) participants required advanced respiratory support with either high-flow nasal cannula, non-invasive positive pressure ventilation, or invasive mechanical ventilation during their hospitalization. Of the remaining participants, 78.0% required low-flow supplemental oxygen and 4.4% did not require oxygen support. The median hospital length of stay was eight (IQR 6-12) days. Of the 40 participants admitted to the ICU, the median ICU length of stay was 7 (IQR 4-15) days.

**Table 3.** Clinical characteristics of VALIANT participants during COVID-19 hospitalization

| Characteristic | VALIANT Participants<br>(N = 341)<sup>a</sup> |
| --- | --- |
| Highest level of care, n (%) |  |
| Floor | 272 (79.8) |
| Stepdown unit | 29 (8.5) |
| Intensive care unit | 40 (11.7) |
| Highest level of respiratory support, n (%) |  |
| None | 15 (4.4) |
| Nasal cannula | 266 (78.0) |
| High flow oxygen | 36 (10.6) |
| Non-invasive positive pressure ventilation | 10 (2.9) |
| Mechanical ventilation | 14 (4.1) |
| Received vasopressors, n (%) | 18 (5.3) |
| SOFA score, maximum (range, 0-24), median (IQR)<sup>b</sup> | 3 (2-4) |
| Hospital length of stay, days, median (IQR) | 8 (6-12) |
| ICU length of stay, days, median (IQR) | 7 (4-15) |
| Died in hospital after the baseline interview, n (%) | 5 (1.5) |
| COVID-specific treatments given, n (%) |  |
| Remdesivir | 246 (72.1) |
| Tocilizumab | 68 (19.9) |
| Corticosteroids<sup>c</sup> | 239 (70.1) |
| Selected laboratory values |  |
| High-sensitivity C-reactive protein (maximum), mg/L, mean (SD) | 109.6 (84.3) |
| D-dimer, mg/L FEU (maximum), median (IQR) | 1.4 (0.8-3.4) |
| Interleukin-6, pg/mL (maximum), median (IQR) | 20.6 (8.9-58.6) |
| Procalcitonin value suggestive of high likelihood of bacterial infection, n (%)<sup>d</sup> | 63 (19.0) |
| Elevated troponin, n (%)<sup>e</sup> | 118 (44.7) |
| Body mass index, mean (SD)<sup>f</sup> | 31.0 (7.2) |
| Number of symptoms endorsed (range, 0-14), mean (SD) | 9 (3) |
| Symptom, n (%) |  |
| Fatigue | 276 (89.3) |
| Weakness | 265 (85.8) |
| Cough | 257 (83.2) |
| Sleepiness (daytime) | 241 (78.0) |
| Loss of appetite | 230 (74.4) |
| Shortness of breath | 212 (68.6) |
| Sleeplessness (night-time) | 200 (64.7) |
| Pain | 178 (57.6) |
| Diarrhea | 169 (55.2) |
| Dizziness | 155 (50.2) |
| Ageusia | 150 (48.5) |
| Trouble thinking | 140 (45.3) |
| Nausea | 120 (38.8) |
| Anosmia | 114 (36.9) |
| Total modified Edmonton Symptom Assessment Score<br>(range, 0-42), mean (SD)<sup>g</sup> | 19 (9) |
| Cognitive impairment, n (%)<sup>h</sup> | 124 (45.3) |
| Any delirium during hospitalization, n (%)<sup>i</sup> | 23 (6.7) |
| Depression over past two weeks<sup>j</sup> | 93 (30.4) |
| Anxiety over past two weeks<sup>k</sup> | 66 (21.5) |
| Physical therapy or occupational therapy ordered, n (%) | 17 (5.0) |
| Code status, n (%)<sup>l</sup> |  |
| Full | 299 (87.7) |
| Limited | 36 (10.6) |
| Comfort measures | 6 (1.8) |
| Transferred from an outside hospital, n (%) | 6 (1.8) |
| Discharge destination |  |
| Home | 176 (51.6) |
| Home with services | 106 (31.1) |
| Skilled nursing facility | 47 (15.0) |
| Long-term acute care facility | 2 (0.6) |
| Inpatient hospice | 2 (0.6) |
| Expired in hospital after enrollment | 5 (1.5) |
| Left against medical advice | 3 (0.9) |

<sup>a</sup> There were missing data for the following variables (number of participants with missing values noted in parentheses): cognitive impairment (67), symptoms during COVID-19 illness (36), depression (35), anxiety (34), high-sensitivity C-reactive protein (12), D-Dimer (1), interleukin-6 (101), procalcitonin (9), troponin (77), and delirium (26).
<sup>b</sup> SOFA = Sequential Organ Failure Assessment. The SOFA score is based on oxygenation, platelet count, Glasgow Coma Scale, bilirubin, mean arterial pressure or receipt of vasoactive agents (dopamine, dobutamine, epinephrine, or norepinephrine), and creatinine. Scores range from 0-24, where higher scores indicate worse severity of illness.
<sup>c</sup> Among participants, 221 received dexamethasone, 10 received hydrocortisone, 18 received methylprednisolone, and 35 received prednisone; 35 participants received more than 1 corticosteroid.
<sup>d</sup> Any procalcitonin value >0.5 ng/mL.
<sup>e</sup> Any troponin value above the upper limit of normal, as reported by the manufacturer of the assay used at each study site.
<sup>f</sup> Body mass index was collected from the index admission or the closest measure within six months prior to index admission in the electronic health record.
<sup>g</sup> The 14 symptoms above were combined into a modified version of the Edmonton Symptom Assessment System scale that accounts for symptom severity (0= symptom absent, 1= mild, 2= moderate, 3= severe). A higher score indicates greater symptom burden.
<sup>h</sup> Assessed through the Montreal Cognitive Assessment 5-minute protocol, with a score ≥22 indicating no cognitive impairment and a score <22 indicating cognitive impairment.
<sup>i</sup> Based on Confusion Assessment Method or Confusion Assessment Method-ICU assessments in the electronic health record.
<sup>j</sup> Assessed through two items on the Patient Health Questionnaire-4, asking how often over the past two weeks the participant felt down, depressed, or hopeless and how often there was little interest or pleasure in doing things. Each item was scored from 0 (not at all) to 3 (nearly every day). A total score ≥3 was interpreted as positive for depression and <3 as negative for depression.
<sup>k</sup> Assessed through two items on the Patient Health Questionnaire-4, asking how often over the past two weeks the participant felt nervous, anxious, or on edge, and how often the participant was unable to stop or control worrying. Each item was scored from 0 (not at all) to 3 (nearly every day). A total score ≥3 was interpreted as positive for anxiety and <3 as negative for anxiety.
<sup>1</sup> Abstracted through manual chart review. “Limited” indicates that there were restrictions on care, such as a do-not-resuscitate or do-not-intubate order. When present, comfort measures orders were placed after study enrollment, since patients with such orders at the time of screening were excluded from participation. For participants whose code status changed during hospitalization, the code status with the most restrictions on care was chosen.

The majority of participants (239, or 70.1%) received a corticosteroid during their hospitalization, which in most cases was dexamethasone. Remdesivir was administered to 246 (72.1%) participants and tocilizumab to 68 (19.9%) participants. Considering the maximum value reported for each study participant during hospitalization, the median D-Dimer was 1.4 (IQR 0.8-3.4) mg/L Fibrinogen Equivalent Units and the median interleukin-6 (IL-6) value was 20.6 (IQR 8.9-58.6) pg/mL, although IL-6 values were missing for 101 patients. During the baseline interview, participants reported a mean of 9 (SD 3) out of 14 possible COVID-19-related symptoms and an average symptom burden of 19 (SD 9), out of a maximum of 42, when each symptom was graded according to severity. There were 124 (45.3%) participants who had a score of <22 on the MoCA 5-minute protocol, suggestive of cognitive impairment. Delirium was present in 23 (6.7%) participants.

## Discussion

We designed and conducted VALIANT to provide detailed, high-quality data about the health outcomes of older adults who have survived a COVID-19 hospitalization. We placed particular emphasis on evaluating the outcomes that matter most to older persons, including preservation of physical function and cognition as well as freedom from burdensome symptoms.^10^ Among the 341 participants we enrolled, there were high rates of baseline functional disability, high rates of cognitive impairment, and high symptom burden. The frequent and detailed assessments performed following hospital discharge will provide a rich opportunity to examine these issues and how they change over the six months after participants left the hospital.

There is a considerable and growing body of literature examining clinical outcomes following COVID-19 infection. Prior research has used administrative data to examine the post-acute sequelae of disease^37^ and to describe functional impairment among COVID-19 survivors who receive home health care services.^38^ There have been studies assessing persistent symptoms following hospitalization,^39, 40^ the development of new disability,^41, 42^ and cognitive impairment,^43^ with investigators in the Post-Hospitalization COVID-19 Study recently using cluster analysis to identify four distinct recovery phenotypes among a large of cohort of patients discharged from a hospital in the United Kingdom.^44^ VALIANT adds to this work in several important ways. First, we specifically focused on older patients, who are much more likely to have serious illness with COVID-19 and are also at much greater risk of functional and cognitive impairment following hospitalization.^45, 46^ The mean age of our cohort, 71.4 years, is substantially higher than those of prior studies. Second, existing cohorts of COVID-19 survivors have nearly all collected data during a single interview.^41, 43, 44^ One exception involves patients in Wuhan, China, who were hospitalized early in the COVID-19 pandemic and then evaluated at 6 and 12 months after infection.^47^ This cohort was considerably younger than ours, however, and participants did not undergo assessment at the time of hospitalization or provide information about function or cognition. VALIANT was designed to overcome these limitations by including a baseline interview, performed during or as close to the acute illness as possible, and by obtaining repeated interviews during the six months following hospital discharge. Since the available evidence suggests that recovery among older persons is highly dynamic, with frequent transitions between states of disability and independence,^48^ this approach will allow us to shed new light on the trajectories of function, cognition, and symptom burden that older adults experience following hospitalization with COVID-19. We expect that these high-quality data will guide the development of interventions to improve the health and well-being of older survivors and to contribute to the evidence base that informs medical decisions for these patients.

Our study has several additional strengths. COVID-19 has caused serious illness disproportionately in historically marginalized racial and ethnic groups that have often been reluctant to engage in medical research.^49^ The racial and ethnic makeup of the VALIANT cohort approximates the racial and ethnic makeup of the two counties from which most participants were recruited.^50^ Specifically, more than 20% of participants were Black and more than 13% were Hispanic. In addition, participants were socioeconomically diverse, with 30% covered by Medicaid, and their baseline health characteristics mirror the heterogeneity of older adults seen in clinical practice. Participants had a median of three chronic conditions, nearly half were disabled in at least one functional activity prior to hospitalization, and a majority were frail or pre-frail. Additionally, there were minimal missing data, except in laboratory values that were eliminated from the standard hospital order set during the study period.

There are two important limitations to these data. The first is that all assessments were conducted remotely. While the study team initially planned to conduct the final, six-month visit in person, this proved infeasible because of restrictions put in place during the pandemic. We instead constructed study interviews with items that could be measured reliably and with high validity via telephone or videoconference. The second is that, while most eligible patients agreed to participate, only slightly more than half of consented patients completed the baseline assessment, despite efforts by our highly experienced and dedicated research staff. Research personnel reported that many patients simply felt too sick during their infection with COVID-19 to participate in a research study. This raises the possibility that our cohort is biased towards patients who were less ill during hospitalization, and, indeed, only 20% of VALIANT participants were admitted to an ICU or stepdown unit. However, nearly all required supplemental oxygen, the majority qualified for corticosteroids and remdesivir, and the median hospital length of stay was eight days — characteristics that reflect the presence of severe illness.

In conclusion, VALIANT is a unique longitudinal study of older adults who have survived hospitalization with COVID-19. It contains granular measures, reported during acute illness and then at multiple time points afterward, that were selected to provide evidence about the health outcomes that matter most to older persons. We anticipate that the knowledge gained from this rich resource will ultimately help guide the development of interventions directed at improving the health of older COVID-19 survivors.

## Author contributions

Study concept and design — Cohen, Hajduk, Ferrante.

Acquisition of subjects and data — Cohen, Geda, Lee, Hajduk, Ferrante.

Analysis and interpretation of data — All authors.

Preparation of manuscript — All authors.

## Other acknowledgments

For help with participant recruitment and interviews, the authors express their gratitude to the members of the Operations Core at the Yale Claude D. Pepper Older Americans Independence Center, including Andrea Benjamin, Sandi Capelli, Amanda Dressel, Kizzy Hernandez-Bigos, Emily Marry, Bridget Mignosa, Amy Shelton, and Maria Zenoni. The authors also wish to thank Harry Doernberg for assistance with chart reviews and the Joint Data and Analytics Team at Yale School of Medicine for assistance with accessing data from the electronic health record. We would like to thank the Yale Department of Medicine COVID-19 Explorer data repository, which was made possible by funding from the Department of Medicine, the George M. O’Brien Kidney Center at Yale (P30DK079310), resources from the Clinical and Translational Research Accelerator, and collaboration with the Yale Center for Clinical Investigation.

