## Supplementary Material for "Rationale, Design, and Baseline Characteristics of the VALIANT (COVID-19 in Older Adults: A Longitudinal Assessment) Cohort"

1. **Supplemental Table 1**: Schedule of assessments in VALIANT
2. **Supplemental Table 2**: Definition and operationalization of selected variables

**Supplemental Table 1.** Schedule of assessments in VALIANT

| Assessment Domain | Interview | | | |
|---|---|---|---|---|
| | **Baseline** | **1-month** | **3-month** | **6-month** |
| Physical function[a] | X | X | X | X |
| Cognitive function | X | X | X | X |
| Symptoms | X | X | X | X |
| Physical activity | X | X | X | X |
| Health status | X | X | X | X |
| Hearing | X | X | X | X |
| Depression | X | X | X | X |
| Anxiety | X | X | X | X |
| Social support | X | X | X | X |
| COVID-19 vaccination status[b] | X | X | X | X |
| Frailty | X | | | X |
| Falls | X | | | X |
| Vision | X | | | |
| Fracture risk | X | | | |
| Clinical characteristics[c] | X | | | |
| Sociodemographic information | X | | | |
| Functional recovery expectations | | X | X | X |
| Post-index admission events | | | | |
| Restricted activity | | X | X | X |
| Fractures | | X | X | X |
| Receipt of PT and OT | | X | X | X |
| Emergency department visits | | X | X | X |
| Hospital readmissions | | X | X | X |
| Admission to a skilled nursing facility | | X | X | X |
| Receipt of home health care | | X | X | X |

*Abbreviations*: PT = physical therapy; OT = occupational therapy.

[a] Information was collected through interviews with the participant or proxy, conducted via telephone or videoconference. Certain modules were omitted in proxy interviews; see Supplementary Table 2.

[b] For assessments performed prior to December 1, 2020, vaccination status was automatically assigned as "no."

[c] Clinical characteristics and sociodemographic information were collected from the Yale Department of Medicine-COVID-19 Data Explorer (DOM-CovX) registry and manual chart abstraction. Because information about delirium was not present in the DOM-Cov-X registry, it was retrieved from the electronic health record via a separate data pull by the Joint Data Analytics Team at the Yale New Haven Health System.

**Supplemental Table 2.** Definition and operationalization of selected variables

| Variable | Definition | Operationalization | Administration | Reference |
|---|---|---|---|---|
| Hearing impairment | Participants asked if:<br>1. They used a hearing aid;<br>2. Could hear well enough to carry on a conversation in a quiet room;<br>3. Could hear well enough to carry on a conversation in a noisy room;<br>4. Their hearing was “excellent,” “good,” “poor,” “very poor,” or if they were deaf. | Hearing impairment present if participant answered “yes” to using a hearing aid, reported they could not hear well enough to carry on a conversation in a quiet or noisy room, or stated that they were deaf. | Participant or Proxy | 24 |
| Vision impairment | Participants asked if they had difficulty (with corrective lenses, as applicable):<br>1. Reading the newspaper<br>2. Recognizing a person from across the street<br>3. Watching television from across the room | Vision impairment present if participant answered “yes” to at least one question. | Participant or Proxy | 24 |
| Disability in functional activities | Participants asked if they needed help from another person to complete each of 15 activities before they became sick with COVID-19:[a]<br>1. Dressing<br>2. Bathing<br>3. Eating<br>4. Personal grooming<br>5. Using the toilet<br>6. Getting in and out of a chair<br>7. Walking around house or apartment<br>8. Going shopping<br>9. Preparing a meal<br>10. Taking medications<br>11. Managing finances<br>12. Doing housework<br>13. Walking a quarter mile<br>14. Climbing up flight of stairs<br>15. Lifting or carrying a 10-pound object | Participant assigned 1 point for each activity requiring assistance, for a total score from 0-15. | Participant or Proxy | 13, 14, 15 |

| | | | | |
|---|---|---|---|---|
| Frailty | Calculated by adding the scores from the five individual Fried frailty items, below. | 0: Not frail<br>1-2: Pre-frail<br>≥3: Frail | Participant or Proxy | 21 |
| Frailty: Low physical activity | Participant asked:<br>1. "In the month before your hospitalization, how often did you take a walk for exercise?"<br>2. "In the month before your hospitalization, did you ever spend time on vigorous activities that increased your heart rate and made you breathe harder? This includes things like working out, swimming, running or biking, or playing a sport."[b] | 0: "Seldom," "sometimes," or "often" to all questions<br>1: "Never" to at least one question | Participant or Proxy | 23 |
| Frailty: Slow walking pace | Participant asked: "How would you describe your usual walking pace?" | 0: Brisk or average pace<br>1: Slow pace or unable to walk | Participant or Proxy | 22 |
| Frailty: Difficulty opening jar | Participant asked: "How much difficulty do you have when opening a jar of jam (or jar of something else) that has never been opened?" | 0: No difficulty or a little difficulty<br>1: Moderate difficulty or a lot of difficulty | Participant or Proxy | 22 |
| Frailty: Unintentional weight loss | Participant asked: "In the last year (12 months) before you had COVID-19, have you lost more than 10 pounds?"[c] | 0: No<br>1: Yes | Participant or Proxy | 21 |
| Frailty: Exhaustion | Participant asked: "For each of the following statements, please tell me how often you felt this way about a month before you were hospitalized:<br>1. I felt that everything I did was an effort.<br>2. I could not get 'going.'"[d] | 0: "Rarely or never" or "some of the time" to all questions<br>1: "Much or most of the time" to at least one question | Participant only | 21 |
| Cognitive impairment | Administered Montreal Cognitive Assessment 5-Minute Protocol | Scores range from 0-30, with scores <22 suggestive of cognitive impairment | Participant only | 16 |

| | | | | |
|---|---|---|---|---|
| Symptom burden | Participants were asked if they had experienced the following symptoms since the onset of their COVID infection:[e]<br><br>1. Fatigue<br>2. Shortness of breath<br>3. Cough<br>4. Loss of smell<br>5. Loss of taste<br>6. Problems with appetite<br>7. Nausea<br>8. Diarrhea<br>9. Daytime sleepiness<br>10. Difficulty sleeping at night<br>11. Pain<br>12. Weakness<br>13. Dizziness<br>14. Difficulty thinking.<br><br>If present, participants were then asked how severe each symptom was at its worst. | Each symptom was scored as follows:<br><br>0: Not present<br>1: Mild severity<br>2: Moderate severity<br>3: Severe severity<br><br>Individual symptom scores were summed into a total symptom burden score, ranging from 0-42. | Participant only | 18, 19 |
| General health | Participant asked: "In general, about a month before you came to the hospital, would you say your health is excellent, very good, good, fair, or poor?"[f] | 1: Excellent<br>2: Very good<br>3: Good<br>4: Fair<br>5: Poor | Participant only | 20 |

| | | | | |
|---|---|---|---|---|
| Depression | Participant asked: "In the past 2 weeks, how often did you feel bothered by any of the following:<br><br>1. Little interest or pleasure in doing things.<br>2. Feeling down, depressed, or hopeless." | Each question scored as follows:<br><br>0: Not at all<br>1: Several days (1-3 days)<br>2: More than half of the days (7-10 days)<br>3: Nearly every day (11-14 days)<br><br>Sum of scores ≥3 indicates positive screen for depression. | Participant only | 26 |
| Anxiety | Participant asked: "In the past 2 weeks, how often did you feel bothered by any of the following:<br><br>1. Feeling nervous, anxious, or on edge.<br>2. Not being able to stop or control worrying." | Each question scored as follows:<br><br>0: Not at all<br>1: Several days (1-3 days)<br>2: More than half of the days (7-10 days)<br>3: Nearly every day (11-14 days)<br><br>Sum of scores ≥3 indicates positive screen for anxiety. | Participant only | 26 |
| Social support | Participant asked: "A month before your hospital stay, how often did you have:<br><br>1. Someone to confide in;<br>2. Someone to help with chores;<br>3. Someone to turn to for suggestions;<br>4. Someone to love and make you feel wanted;<br>5. Someone to relax with."[g] | Items scored from 1-5,<br><br>1: None of the time<br>2: A little of the time<br>3: Some of the time<br>4: Most of the time<br>5: All of the time<br><br>Scores then summed for total score ranging from 5-25. | Participant only | 27 |

| | | | | |
|---|---|---|---|---|
| History of falls | Participant asked: "In the last year (12 months) before you had COVID, have you had a fall?"[h] | 0: No self-reported fall<br>1: ≥1 self-reported fall | Participant or proxy | N/A |
| History of fractures | Participant asked: "Have you broken a bone since you turned 50 years old?" | 0: No self-reported fractures<br>1: ≥1 self-reported fracture | Participant or proxy | 30 |

[a] At follow-up interviews, participants or proxies were asked about whether the need for help at the present time.

[b] At the one month follow-up interview, participants or proxies were asked about physical activity in the prior month; at the three month follow-up, they were asked about physical activity in the prior two months; at the six month follow-up interview, they were asked about physical activity in the prior three months.

[c] At the six month follow-up interview, participants or proxies were asked about weight loss since diagnosis with COVID-19.

[d] At follow-up interviews, participants were asked about exhaustion in the past two weeks.

[e] At follow-up interviews, participants were asked about symptoms experienced within the past 24 hours.

[f] At follow-up interviews, participants were asked about their health at the present time.

[g] At follow-up interviews, participants were asked about social support in the past month.

[h] At the six month follow-up interview, participants or proxies were asked about falls since diagnosis with COVID-19.
